# Performance evaluation of the Xpert MTB/XDR test for the detection of drug resistance to *Mycobacterium tuberculosis* among people diagnosed with tuberculosis in South Africa

**DOI:** 10.1101/2024.02.16.24302824

**Authors:** Shaheed Vally Omar, Gail Louw, Farzana Ismail, Xiaohong Liu, Dumsani Ngcamu, Thabisile Gwala, Minty van der Meulen, Lavania Joseph

**Affiliations:** Centre for Tuberculosis, National TB Reference Laboratory, National Institute for Communicable Diseases a division of the National Health Laboratory Service, Johannesburg, South Africa; Cepheid (PTY) LTD, Johannesburg, South Africa; Cepheid, Sunnyvale, California, USA

**Keywords:** drug-resistant, TB, molecular, XDR, diagnostic

## Abstract

**Background:** Tuberculosis (TB) remains a significant public health concern in South Africa, with high incidence rates and a growing burden of drug-resistant TB. This study aimed to assess the diagnostic performance of the Xpert MTB/XDR test, a novel molecular assay, for detecting drug resistance in TB patients using archived sputum sediments.

**Methods:** The study involved a comprehensive analysis of 322 samples collected from presumptive TB patients between 2016 - 2019 across South Africa, previously characterized by phenotypic and genotypic methods. The Xpert MTB/XDR test was evaluated for its ability to detect resistance to isoniazid (INH), ethionamide (ETH), fluoroquinolones (FLQ), and second-line injectable drugs (SLID) compared to phenotypic drug susceptibility testing (pDST) and whole-genome sequencing (WGS). The Xpert MTB/RIF Ultra and G4 tests were performed to determine agreement with this test for TB detection.

**Findings:** The Xpert MTB/XDR test performance showed excellent sensitivity and specificity for detecting Mycobacterium tuberculosis (*M. tuberculosis*), with a sensitivity of 98.3% and specificity of 100% compared to culture. The sensitivities using a composite reference standard, pDST and sequencing, were over 90% for INH, FLQ, AMK, KAN, and CAP resistance, meeting the WHO target product profile criteria for this class. A lower sensitivity of 65.9% for ETH resistance was observed, driven by the limited targets covered by the assay.

**Interpretation:** The Xpert MTB/XDR test offers a promising solution for the rapid detection of drug-resistant TB in South Africa. It could significantly enhance TB control efforts in this setting and contribute to improved patient care and management.

## Introduction

Progress in achieving the 2018 UN High-Level Meeting targets has been exceptionally poor for multidrug-resistant (MDR)/Rifampicin Resistant (RR) tuberculosis (TB) with only 55% of the target achieved between 2018-2022 (1). In a recent commentary on diagnostics for TB, the authors succinctly summarized the situation as “… if we cannot find TB, we cannot treat TB. And if we cannot treat TB, we cannot end TB”, highlighting the central role of diagnostics to address the gaps (2).

Over the past decade, South Africa has made significant progress towards the prevention and management of TB disease and drug-resistant TB since adopting the WHO’s recommended diagnostic technologies and therapies. South Africa is now one of six high-burden countries that achieved the 2020 End TB Strategy milestones of a 20% reduction in TB incidence rates (3) and has sustained this milestone in 2021 (4). It is ranked as one of the WHO’s top 30 high-burden countries MDR/RR-TB (4) and has a disproportionate burden on the African continent, with 32% (6781/21402) of the laboratory-confirmed cases reported from this country alone (1).

One of the key elements for meeting the End TB strategy (5), is the recommendation to use WHO-endorsed rapid molecular tests, such as the Xpert^®^ MTB/RIF (“G4”) (Cepheid, Sunnyvale, CA, USA) for the diagnosis of TB and the detection of rifampicin (RIF) resistance in patients presumptive of TB rather than smear microscopy (6) and was fully implemented in 2013 with a subsequent transition to Xpert^®^ MTB/RIF Ultra (“Ultra”) in 2017.

Another target set in the WHO End TB Strategy is for all MDR/RR-TB patients to be tested for fluoroquinolone (FQ) resistance. In 2022, only 50% of all MDR/RR-TB patients were tested globally (1). The Bruker-Hain GenoType MTBDR*plus* and GenoType MTBDR*sl* line probe assays (LPAs) and phenotypic drug susceptibility test (DST) based where genotypic methods are not available have been implemented nationally. However, these assays require specialized facilities and it can take between 1-4 weeks for results to be available (7, 8).

An important priority highlighted by WHO has been for a TB DST that is fast, with low technical skill and minimal infrastructure requirements (9). In response, Cepheid announced the release of the Xpert^®^ MTB/XDR test in July 2020, a test capable of detecting resistance using melt curve analysis by targeting associated mutations to isoniazid (INH), ethionamide (ETH), fluoroquinolones (FLQ), and second-line injectable agents (SLID), such as kanamycin (KAN), amikacin (AMK) and capreomycin (CAP) (10). The WHO have since endorsed the MTB/XDR test as a low complexity molecular sputum-based reflex test for the detection of resistance to these drugs in confirmed *M. tuberculosis* positive specimens (6). The test targets resistance conferring mutations in the *katG* and *fabG1* genes, *ahpC-oxyR* intergenic region and *inhA* promoter for INH; *inhA* promoter mutations only for ETH resistance; the *gyrA* and *gyrB* quinolone resistance determining regions (QRDR) for FLQ; and the *rrs* gene and the *eis* promoter region for SLID. It reports low-level resistance to INH and FLQ and also calls resistance to select SLID base on mutations detected.

The main study that assessed the clinical performance of the Xpert MTB/XDR test was conducted at 2 study sites (South Africa and China), however the data presented in this manuscript is focused on the South Africa data set only.

This study aimed to evaluate the performance of the Xpert MTB/XDR test to detect drug resistance using phenotypic DST and whole genome sequencing (WGS) restricted to test targets as the reference standard and success rate of the assay compared to the G4 and Ultra test.

## Materials and Methods

### Ethics

The study was conducted at the Centre for Tuberculosis, National TB Reference Laboratory at the National Institute for Communicable Diseases a division of the National Health Laboratory Service in Johannesburg, South Africa. Ethical approval M160667 for this study was obtained from the University of the Witwatersrand, Johannesburg Human Research Ethics Committee.

### Sample Eligibility Criteria

Archived sputum sediments, prepared from sputum specimens collected between 29 November 2016 to 20 June 2019 from people with presumptive TB, representative of all nine provinces in South Africa, were used in this study. These samples were characterized by phenotypic and genotypic methods and stored at −70°C.

*M. tuberculosis* positive and *M. tuberculosis* negative specimens were included for evaluation if a minimum volume of 1mL frozen sputum sediment was available for testing (11). For *M. tuberculosis* positive specimens, documented results for smear microscopy, positive culture result on the BD BACTEC Mycobacterial Growth Indicator Tube 960 (MGIT) platform (BD Biosciences, Sparks, MD, USA) and/or Lowenstein Jensen (LJ) media were required. In addition, pDST results for isoniazid, ethionamide, amikacin, kanamycin, capreomycin and moxifloxacin/levofloxacin on the MGIT platform; sequencing results for *katG, inhA* promoter, *fabG1*, *oxyR-ahpC* intergenic region, *gyrA*, *gyrB*, *rrs* and *eis* promoter; and G4 or Ultra test results were also required.

For *M. tuberculosis* negative specimens, documented results indicating negative culture result on MGIT and/or LJ media was required. Specimens that were previously thawed were excluded from the study.

### Specimen Processing Procedures

Concentrated sediments were prepared from induced or expectorated sputum and subsequently stored at −70°C. These specimens were assigned a unique de-identified specimen ID by a third-party individual not actively involved in the study. Subsequently, the specimens were decontaminated using a commercial NaLC/NaOH kit, Alphatec NAC-PAC™ Red (Alpha-Tec Systems, Inc, Vancouver, WA, USA), with a final concentration of 1.5% sodium hydroxide as per manufactures instruction. Two drops of the sediment were used to prepare a smear for Auramine-O staining and were graded in accordance with Global Laboratory Initiative Guidelines (12).

A 500µL aliquot of the sediment was inoculated into a MGIT tube and incubated at 37°C in the MGIT 960 system (BD Biosciences, Sparks, MD, USA), until the culture flagged positive or 42 days to a negative result as per the manufacturers instruction. The MGIT positive cultures were confirmed to be MTB complex by antigen testing (TBCheck, BD, Sparks, USA) and were inspected for contamination by visual observations, Ziehl Neelsen (ZN) staining and on blood agar prior to performing pDST.

### Reference method testing (pDST and WGS)

The MGIT 960 platform and BD Epicentre TBeXiST module (BD Biosciences, Sparks, MD, USA) was used to perform pDST as previously described (13) which included, INH, ETH, FLQ (moxifloxacin(MXF), levofloxacin (LEV) and ofloxacin (OFL)) and SLID (AMK, KAN and CAP). Testing was performed for all MTB/XDR drug targets using the 2018 updated WHO recommended critical concentrations (14). Whole genome sequencing (WGS) and variant analysis was performed as previously described (15).

### Specimen testing

#### Xpert MTB/RIF; Xpert MTB/RIF ULTRA and Xpert MTB/XDR test

Sediments were thawed at room temperature prior to testing and were randomly assigned for processing on either the G4 or Ultra tests using the GeneXpert 6-colour module system (Cepheid, Sunnyvale, USA). All sediments were tested on the MTB/XDR test on the GeneXpert 10-colour module system.

Briefly, a 1,2mL aliquot of sediment was transferred to a 15mL conical screw-cap tube and the sample reagent (SR) was added at a 1:3 ratio. The SR-sample mix was homogenized at room temperature by vigorous shaking 10-20 times and incubated for 10 minutes, followed by vigorous shaking for 10-20 times before further incubation for 5 minutes. The SR-sample mix was then used to load either the G4 or Ultra tests and the MTB/XDR test in accordance with the manufacturer’s instructions using the appropriate GeneXpert system. If an “ERROR”, “INVALID” or “NO RESULT” result was obtained by specimen testing, the specimen was re-tested once if sufficient specimen was available. If the re-test result was also non-determinant, the specimen was reported as such.

### Discrepant result resolution

Discrepant results were defined as specimens where the MTB/XDR test and either pDST or sequencing results were not in agreement. These discrepant results were categorized as either a false positive result, which indicates a positive MTB/XDR test result with a negative pDST/or sequencing result or a false negative result, which indicates a negative MTB/XDR test result with a positive pDST/ sequencing result. Specimens with discrepant results were investigated as follows: a) the specimen was re-tested on the Xpert MTB/XDR test, if sufficient volume remained; b) the specimen was tested using either sequencing or pDST that detected the targets that were not in agreement; c) standard of care data associated with the specimen were reviewed to determine the absence or presence of target mutations associated with specific drug resistance and d) the site confirmed that the data from source document matched the data captured electronically.

### Performance Analysis

Clinical performance of MTB/XDR test was evaluated using the following outcome measures: 1) the positive percentage agreement (PPA) and negative percentage agreement (NPA) for *M. tuberculosis* detection against G4 or Ultra, 2) sensitivity and specificity for *M. tuberculosis* detection using MGIT culture as the reference standard; and 3) sensitivity and specificity compared to pDST (for FLQ, the final result was generated considering all drugs tested for this evaluation) and sequencing as reference standard independently and in combination as a composite reference for resistance detection of INH, ETH, FLQs and SLID. The composite reference test results were categorized as “Resistant” if either pDST or Sequencing results were “Resistant”; “Susceptible” if both pDST and sequencing were “Susceptible”. Analysis was performed using RStudio Team (2020) (RStudio: Integrated Development for R. RStudio, PBC, Boston, MA) and the 95% confidence intervals were determined using the Wilson score methods.

We also assessed the ability of the MTB/XDR test to: 1) identify low levels of resistance to INH and FLQ (MXF) based on the detection of resistance causing mutations which typically result in borderline minimal inhibitory concentration (MIC) values that are close to the breakpoint drug concentration used in phenotypic resistance testing; 2) identify resistance to KAN only, KAN and AMI, or cross- resistance to all three SLID and, 3) identify non-determinate rates. Non-determinate test results were excluded from the performance analysis.

## Results

### Specimen characteristics

A total of 324 specimens were eligible for inclusion, of these 2 were excluded due to insufficient specimen volume (Figure 1). The final data analysis was done on 322 specimens which comprised of 46.0% females, 50.3% males and 3.7% of the specimens with unknown sex (Table 1). Additionally, the median age was 37 years (13 – 85) among individuals who had specimens included (Table 1). As shown in Table 1, a positive MGIT culture result was obtained in 92.2% (297/322) of specimens with 74.4% smear positive. A total of 100 specimens were tested on the G4 test, 86.0% (86/100) were MTB positive and 74.4% (64/86) of these were RIF resistant (Table 1, Figure 1). Additionally, 222 specimens were tested on the Ultra test, with a positive result obtained for 91.9% (204/222) of which 30.4% (62/204) were RIF resistant, 68.6% (140/204) RIF sensitive and 1% (2/204) RIF indeterminate (Table 1, Figure 1). An MTB Trace detected result was obtained in 1.8% (4/222) of specimens tested on the Ultra test (Table 1, Figure 1). Overall, 8.7% (28/322) of the specimens were negative by the G4 and Ultra tests (Table 1, Figure 1).

**Figure 1.**
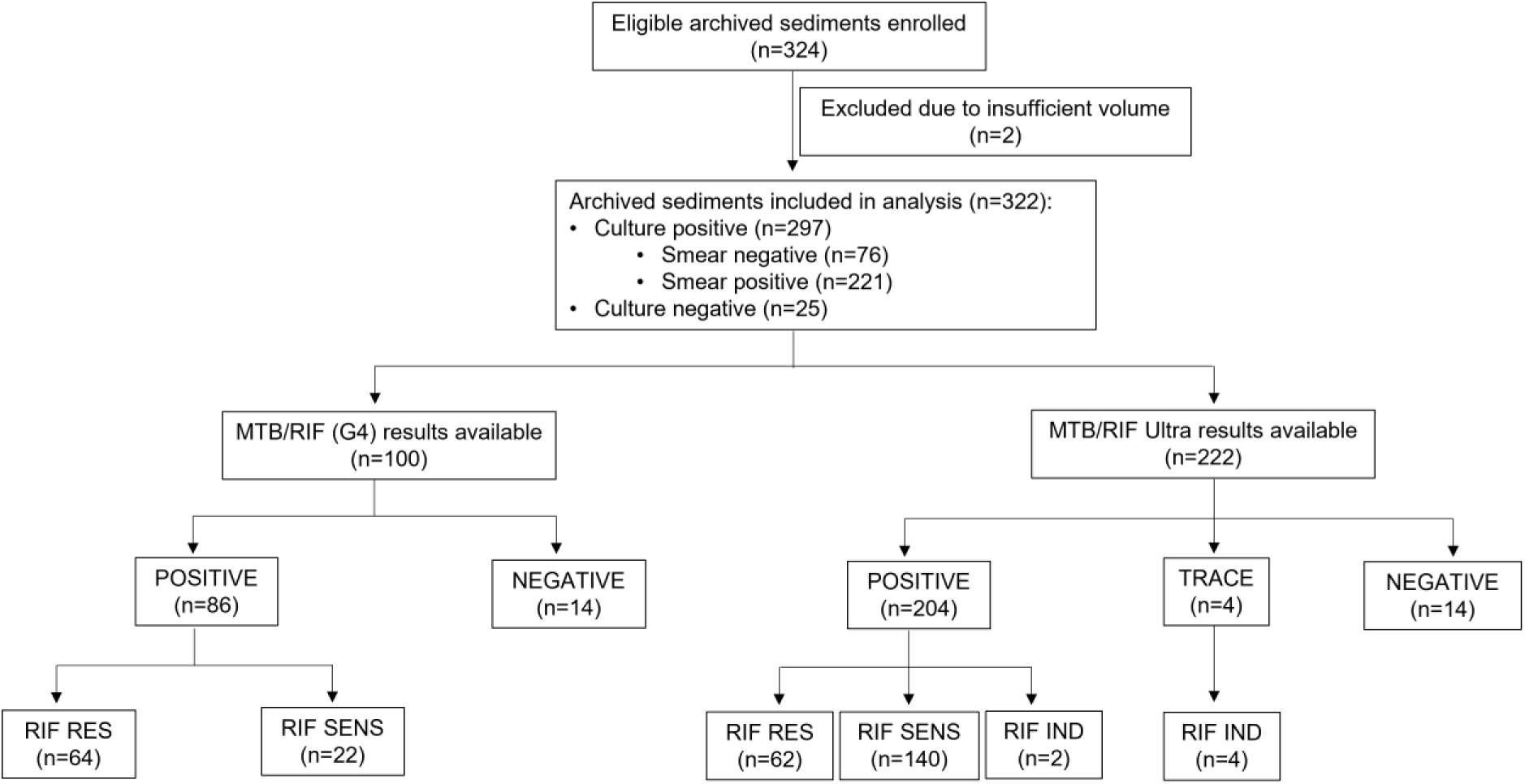
Flowchart illustrating the diagnostic characteristics of the sediments. MTB/RIF POSITIVE constitutes MTB Detected; MTB/RIF Ultra POSITIVE constitutes MTB Detected Very Low, MTB Detected Low, MTB Detected Medium, MTB Detected High; MTB/RIF Ultra TRACE constitutes MTB Trace Detected; MTB/RIF or MTB/RIF Ultra NEGATIVE constitutes MTB Not Detected; RIF RES indicates Rifampicin Resistant, RIF SENS indicates Rifampicin Sensitive, RIF IND indicates Rifampicin Indeterminate.

**Table 1:** Demographics and diagnostic characteristics of specimens included in the analysis.

|  | **Overall**<br>**(N=322)** |
| --- | --- |
| **Demographic Characteristics** |  |
| **Sex** |  |
| Female | 148 (46.0%) |
| Male | 162 (50.3%) |
| N/A | 12 (3.7%) |
| **Age (Years)** |  |
| Median [Min, Max) | 37 (13-85) |
| **MGIT culture / AFB smear** |  |
| % Culture positive [n/N] | 92.2% [297/322] |
| % smear positive [n/N] | 74.4% [221/297] |
| % smear negative [n/N] | 25.6% [76/297] |
| % Culture negative [n/N] | 7.8% [25/322] |
| **G4** |  |
| %G4 results [n/N] | 31.1% [100/322] |
| % Positive<sup>a</sup> [n/N] | 86.0% [86/100] |
| % RIF RES<sup>d</sup> [n/N] | 74.4% [64/86] |
| % RIF SENS<sup>e</sup> [n/N] | 25.6% [22/86] |
| % Negative<sup>c</sup> [n/N] | 14.0% [14/100] |
| **Ultra** |  |
| % Ultra results [n/N] | 68.9% [222/322] |
| % Positive<sup>b</sup> [n/N] | 91.9% [204/222] |
| % RIF RES<sup>e</sup> [n/N] | 30.4% [62/204] |
| % RIF SENS<sup>f</sup> [n/N] | 68.6% [140/204] |
| % RIF IND<sup>g</sup> [n/N] | 1.0% [2/204] |
| % Trace<sup>d</sup> [n/N] | 1.8% [4/222] |
| RIF IND<sup>g</sup> [n/N] | 100% [4/4] |
| % Negative<sup>c</sup> [n/N] | 6.3% [14/222] |

<sup>a</sup>G4 Positive constitutes MTB Detected
<sup>b</sup>Ultra Positive constitutes MTB Detected Very Low, MTB Detected Low, MTB Detected Medium, MTB Detected High
<sup>c</sup>G4 or Ultra Negative constitutes MTB Not Detected
<sup>d</sup>Ultra Trace constitutes MTB Trace Detected
<sup>e</sup>RIF RES indicates Rifampicin Resistant
<sup>f</sup>RIF SENS indicates Rifampicin Sensitive
<sup>g</sup>RIF IND indicates Rifampicin Indeterminate

### Diagnostic performance of MTB/XDR test for the detection of *M. tuberculosis*

The sensitivity of the MTB/XDR test for the detection of *M. tuberculosis* in smear positive, smear negative and overall was 99.5% (95%CI: 97.5 - 99.9), 94.7% (95%CI:87.2 - 97.9) and 98.3% (95%CI: 96.1, 99.3). The specificity was 100% (95% CI: 86.7 – 100) (Table 2).

**Table 2:** Performance summary of the MTB/XDR test relative to MGIT culture, G4 and Ultra test for *M. tuberculosis* detection.

|  | Sensitivity/PPA (95% CI) | Specificity/NPA (95% CI) |
| --- | --- | --- |
| **MGIT Culture<sup>a</sup>** |  |  |
| Overall | 98.3% (96.1 – 99.3) | 100% (86.7 – 100) |
| Culture positive, Smear positive | 99.5% (97.5 – 99.9) | N/A |
| Culture positive, Smear negative | 94.7% (87.2 – 97.9) | N/A |
| **G4<sup>b</sup>** | 98.8% (95%CI: 93.7-99.8) | 100.0% (95%CI: 78.5-100.0) |
| **Ultra<sup>b</sup>** | 99.5% (95%CI: 97.3-99.9) | 100.0% (95%CI: 78.5-100.0) |

<sup>a</sup> Diagnostic Performance of MTB/XDR test in *M. tuberculosis* detection relative to MGIT culture was assessed by sensitivity and specificity.
<sup>b</sup> Diagnostic performance of MTB/XDR test in *M. tuberculosis* detection relative to G4 and Ultra was assessed by PPA and NPA.

The PPA and NPA for MTB/XDR in *M. tuberculosis* detection relative to the G4 test were 98.8% (95%CI: 93.7-99.8) and 100.0% (95%CI: 78.5-100.0), respectively (Table 2). In this instance, the MTB/XDR test failed to detect one sample, with a scant microscopy grading, identified as positive (i.e. MTB detected) by the G4 test. Similarly, the PPA and NPA for MTB/XDR in *M. tuberculosis* detection relative to the Ultra test was 99.5% (95%CI: 97.3-99.9) and 100.0% (95%CI: 78.5-100.0), respectively (Table 2). A smear negative sample identified as MTB Trace Detected by the Ultra test was not detected by the MTB/XDR test.

### Non-determinate rate

The initial non-determinate rate (ND) observed for the MTB/XDR test was 3.1% (10/322) (Data not shown). Of these 0.6% were due to “No Result” and 2.5% due to “Error” results obtained by the MTB/XDR test. Repeat testing of these specimens rendered a valid result with final ND rate of 0% (Data not shown).

### Diagnostic performance of the MTB/XDR test for drug resistance prediction

The diagnostic performance of the MTB/XDR test against pDST, sequencing and the composite reference test for all drugs are shown in Figure 2, 3 and 4, respectively.

**Figure 2:**
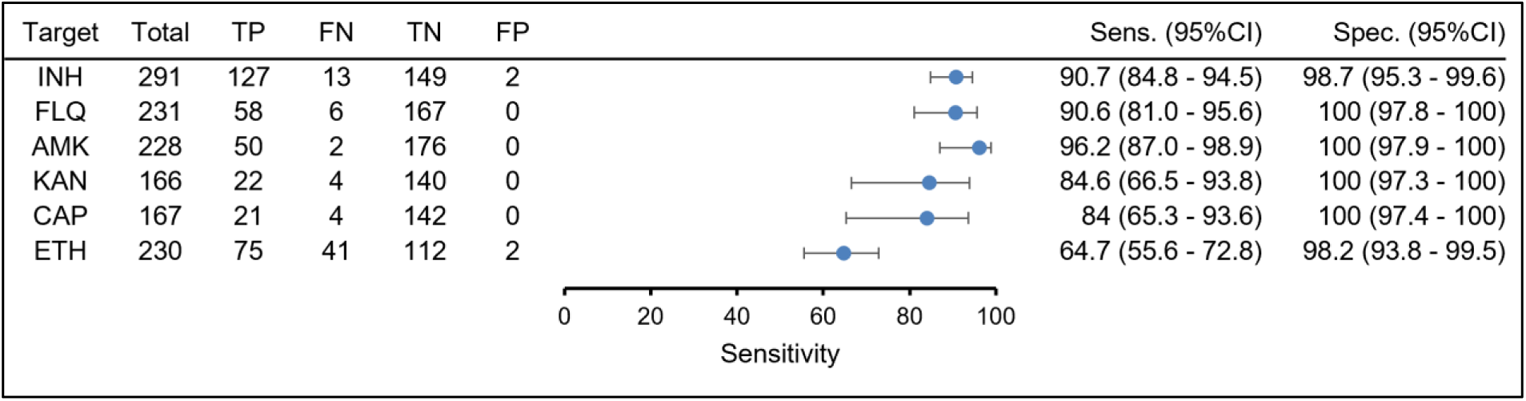
Performance of the MTB/XDR test in predicting resistance to INH, FLQ, AMK, KAN, CAP, ETH relative to pDST. TP - True Positive, FN – False Negative; TN – True Negative; FP – False Positive; Sens. – Sensitivity; Spec. - Specificity

**Figure 3:**
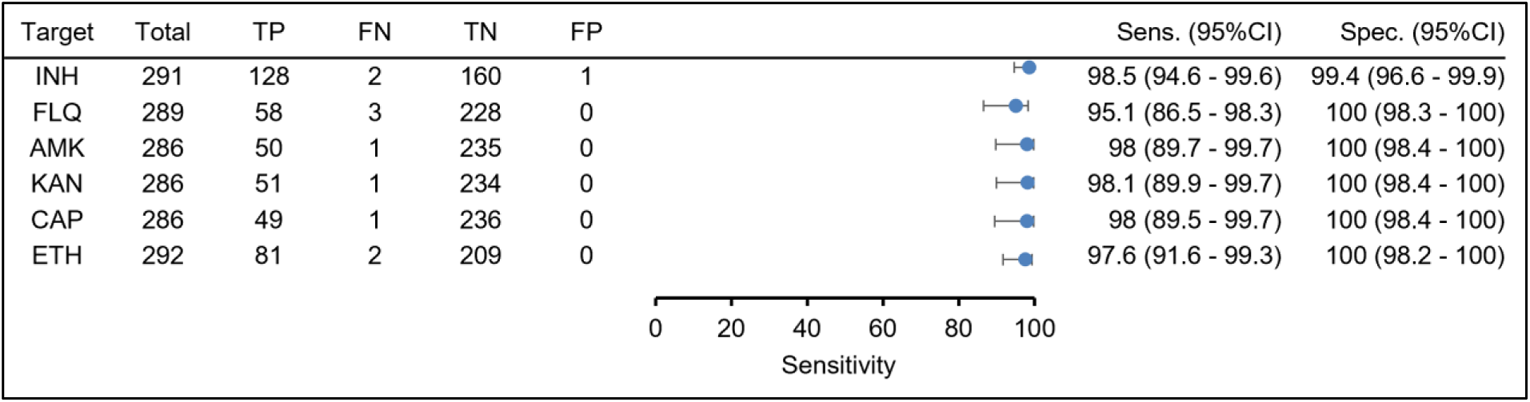
Performance of the MTB/XDR test in detection of resistance to INH, FLQ, AMK, KAN, CAP, ETH relative to sequencing. TP - True Positive, FN – False Negative; TN – True Negative; FP – False Positive; Sens. – Sensitivity; Spec. - Specificity

**Figure 4:**
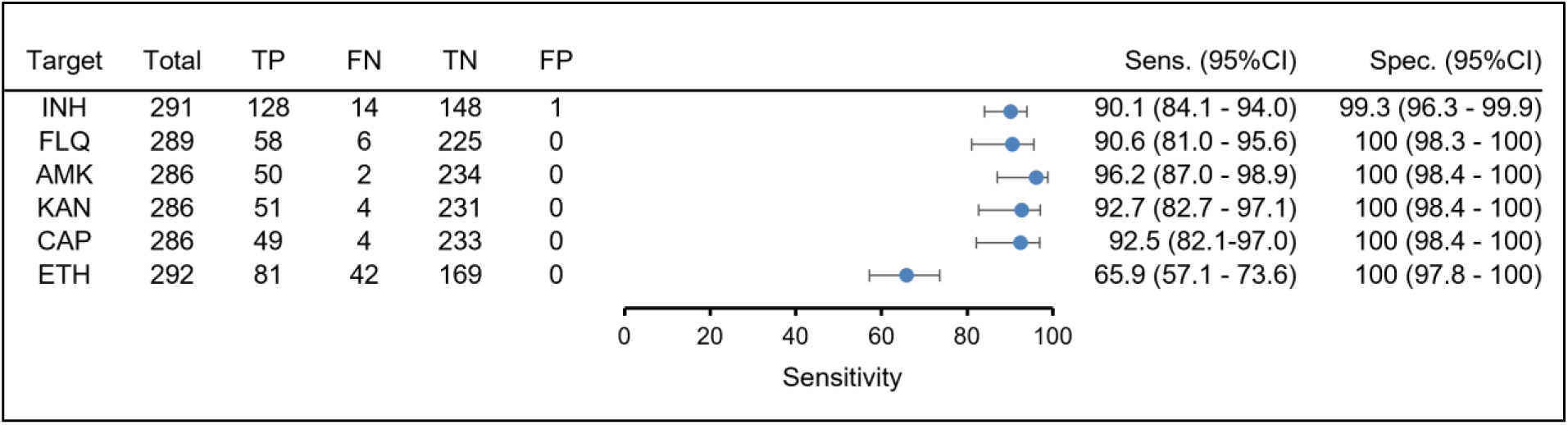
Performance of the MTB/XDR test in detection of resistance to INH, FLQ, AMK, KAN, CAP, ETH relative to the composite test. TP - True Positive, FN – False Negative; TN – True Negative; FP – False Positive; Sens. – Sensitivity; Spec. - Specificity

#### Isoniazid & Ethionamide

Results for the test and reference standard for INH were available for 291 samples. The sensitivity and specificity to detect INH resistance compared to pDST was 90.7% (95% CI: 84.8 – 94.5) and 98.7% (95% CI: 95.3 - 99.6), respectively (Figure 2) while compared to WGS was 98.5% (95% CI: 94.6 – 99.6) and specificity of 99.4% (95% CI: 96.6 – 99.9) respectively (Figure 3). Using a composite reference, the sensitivity was 90.1% (95% CI: 84.1 – 94.0) and a specificity of 99.3% (95% CI: 96.3 – 99.9) (Figure 4).

The prediction of low level of INH resistance using sequencing as reference test was 100% (Supplementary Table 2). The majority (95.8%; 23/24) of samples with low level INH resistance detected harbored the C-15T mutation in the *inhA* promoter and 4.2% (1/24) samples showed the T-8A mutation in the *inhA* promoter (Supplementary Table 2).

Ethionamide (ETH) resistance prediction on the MTB/XDR test is based on detection of mutations in the *inhA* promoter. The sensitivity and specificity in detecting ETH resistance using pDST as the reference was 64.7% (95% CI: 55.6 – 72.8) and 98.2% (95%CI: 93.8 – 99.5) respectively (Figure 2). Using WGS and limited to analysis of the *inhA* promoter only the sensitivity was 97.6% (95% CI: 91.6 – 99.3) (Figure 3). Using the composite reference, the sensitivity was 65.9% (95% CI: 57.1 – 73.6) and specificity of 100% (Figure 3; Figure 4).

#### Fluoroquinolones (FLQs)

A total of 231 samples had available results for both the test and pDST for FLQ and the sensitivity was 90.6% (95% CI: 81.0 – 95.6) (Figure 2). A total of 289 samples had available results for both the test and WGS and the sensitivity was 95.1% (95% CI: 86.5 – 98.3) (Figure 3). Using a composite reference, the sensitivity was 90.6% (95% CI: 81.0 – 95.6) (Figure 4). The specificity was 100% against pDST, WGS and the composite reference (Figure 2, Figure 3, Figure 4).

The ability of the MTB/XDR test to accurately predict low level of FLQ resistance using sequencing as the reference test was 100% (Supplementary Table 3). The distribution of these *gyrA* mutations were Ala90Val (66.7%; 10/15), Asp94Ala (20%; 3/15) and Ser91Pro (13.3%; 2/15) (Supplementary Table 3).

#### Second-line Injectable drugs (AMK, KAN, CAP)

A total of 228 samples had available results for both MTB/XDR and AMK pDST while it was lower for KAN (n=166) and CAP (n=167). The sensitivity in detecting resistance to AMK, KAN and CAP using pDST as the reference were 96.2% (95%CI: 87.0 – 98.9); 84.6% (95%CI: 66.5 – 99.3) and 84.0 (95%CI: 65.3 - 93.6), respectively (Figure 2). The respective sensitivities were higher using WGS as the reference: 98.0% (95%CI: 89.7 - 99.7) for AMK, 98.1% (95%CI: 89.9 - −99.7) for KAN and 98.0% (95%CI: 89.5 - −99.7) for CAP (Figure 3). Using the composite reference, the sensitivity was 96.2% (95%CI: 87.0 – 98.9) for AMK, 92.7% (95%CI: 82.7 – 97.1) for KAN and 92.5% (95% CI:82.1 – 97.0) for CAP (Figure 4). All SLIDs had a specificity of 100% for pDST, sequencing and the composite reference test.

The ability of the MTB/XDR test to accurately predict resistance to the different SLIDs were 100% for each of the anti-TB drugs, with cross-resistance between the SLIDs predicted by the A1401G mutation in the *rrs* gene (Data not shown). Additionally, resistance to KAN only was predicted by the C-12T mutation in the *eis* promoter with resistance to both KAN and AMK predicted by the C-14T mutation in the *eis* promoter (Data not shown).

### Discrepant result resolution

A total of 74 discrepant results were identified between the MTB/XDR test and pDST, with 70 false negative and 4 false positive results identified across all drugs tested (Figure 2).

Four out of four (100%) of the false positive results were resolved by WGS that identified mutations predicting INH and ETH resistance, indicating that the initial susceptible pDST result for both INH and ETH were incorrect.

Sixty-four out of seventy false negative results (91.4%) were resolved by WGS that showed the absence of target gene mutations predicting resistance to INH, FLQ, SLIDs and ETH, indicating that the initial resistant pDST result obtained for the drug targets were likely incorrect in these specimens. The false negative results in six specimens (8.6%) were not resolved by WGS, and the cause of the discrepancy was further investigated. One specimen that was identified as INH resistant by pDST, but INH susceptible by the MTB/XDR test, showed the presence of a Leu203Leu *fabG1* mutation that was not identified by the MTB/XDR test. Three specimens that were identified as FLQ resistant by pDST, but FLQ susceptible by the MTB/XDR test showed the presence of a Asp94Gly (n=2) and Ala90Val mutation (n=1) in the *gyrA* gene. One specimen that was identified as KAN, AMK and CAP resistant by pDST, but susceptible by MTB/XDR showed the presence of a A1401G mutation in the *rrs* gene. Further investigation confirmed heteroresistance, however, the *rrs* mutant melt peak was below the detection threshold of the MTB/XDR test and therefore reported as KAN, AMK and CAP susceptible. Lastly, one specimen identified by the MTB/XDR test as ETH susceptible, but resistant by pDST, had a G-17T mutation in the *inhA* promoter region, which was initially missed by the MTB/XDR test.

A total of 11 discrepant results were identified between the MTB/XDR test and sequencing, with 10 false negative results and 1 false positive result identified (Figure 3). One false positive result was identified with the MTB/XDR test reporting an INH resistant result and sequencing showing an INH susceptible result, although this specimen showed a C-52A mutation in the *ahpC* region, which was not considered associated with INH resistance by the study site.

Out of the 10 false negative results, one specimen was identified as INH susceptible by MTB/XDR test but had a *fabG1* mutation identified by sequencing. Further investigation failed to identify a root cause of the discrepancy, thereby indicating that the MTB/XDR test incorrectly reported this specimen as INH susceptible. The second specimen was incorrectly processed, and no INH resistance causing mutation was identified upon resequencing. Three specimens, identified by the MTB/XDR test as FLQ susceptible, was identified as FLQ resistant by sequencing with the detection of Asp94Gly (n=2) and both Asp94Gly and Asp94Asn (n=1) mutations in the *gyrA* gene. The *gyrA* double mutation of Asp94Gly and Asp94Asn was below the detection threshold of the MTB/XDR test due to hetero-resistant populations and was therefore not identified as FLQ resistance by the test. AMK, KAN and CAP resistance was not detected by the MTB/XDR test in one specimen since the A1401G *rrs* mutant peak was below the detection threshold of the test. Lastly, ETH resistance was not detected by the MTB/XDR test in one specimen due to a mixed culture and in a second specimen due to a specimen swop that showed no *inhA* promoter mutation upon re-sequencing.

## Discussion

The demonstrated diagnostic performance of the MTB/XDR test for *M. tuberculosis* detection met the overall sensitivity (≥80%) and specificity (≥98%) requirements relative to culture as outlined in the guidance for next-generation DST by the WHO (16), with a 98.3% sensitivity and specificity of 100% specificity observed in the current study. The diagnostic performance of the MTB/XDR test was comparable to Ultra (pooled sensitivity 90.9% (95%CI: 86.2 – 94.7)); pooled specificity 95.6% (95%CI: 93.0 – 97.4)) and G4 test (pooled sensitivity of 84.7% (78.6 – 89.9)) and pooled specificity of 98.4% (95%CI: 97.0 – 99.3) as previously described (17). This was also seen in the current study with high levels of agreement between the MTB/XDR and the G4 and Ultra. This can be attributed to the similar limit of detection (LoD) of the MTB/XDR (71.9 CFU/mL (95%CI: 58 to 100)) compared to the G4 (86.9 CFU/mL (95%CI (72 to 110)) (10). The LoD of Ultra (15.6 CFU/mL), however, is lower than that of MTB/XDR (11). Interestingly, the diagnostic sensitivity of the MTB/XDR test in culture positive, smear negative specimens (94.7%) was better than in Ultra (pooled sensitivity of 77.5% (95%CI: 67.6 to 85.6)) and G4 (pooled sensitivity of 60.6% (95%CI: 48.4 to 71.7)) as previously described (17). This indicates that although the MTB/XDR test is a reflex test to be used in *M. tuberculosis* positive cases (excluding ‘Trace’), its ability to accurately detect *M. tuberculosis* in sputum specimens is on par with Ultra and G4. Further studies on processing specimens with MTB ‘Trace’ detected by Ultra which are expected to be lower than the LoD of MTB/XDR and are for reflex testing is required. The MTB/XDR test was able to successfully detect more than 98% of MTB samples detected by the G4 (98.8%) and ULTRA (99.5%) tests, with each comparator test failing to detect one sample each. These were both smear negative and in the case of ULTRA a trace result. These finding are supportive of using the MTB/XDR test as a reflex using the MTB/RIF tests as an indicator for the follow-on testing.

The clinical performance of the MTB/XDR test performance demonstrated by the current study is similar to previous studies (18, 19) with sensitivity of >84% observed for INH, SLIDs and FLQ relative to pDST. The sensitivity for predicting resistance to INH, FLQ and AMK also met the minimum requirement of the target product profile for next-generation pDST (sensitivity of >90% for INH, FLQ and sensitivity of ≥80% for AMK) as specified by the WHO (9, 16). In contrast, lower sensitivity of the MTB XDR test in predicting ETH resistance (sensitivity of 64.7% (95%CI: 55.6 – 72.8)) compared to pDST was demonstrated in the current study, with sensitivity estimates comparable to previously published studies (18, 19). This is due to the inclusion of only the *inhA* promoter target as a proxy for ETH resistance in the MTB/XDR test, since mutations in additional targets (e.g. *ethA*) have been identified to confer resistance to ETH in clinical specimen from TB patients (20, 21).

When considering sequencing alone, the clinical performance of the MTB/XDR assay for all drug targets increased to >95% sensitivity and >99% specificity (Figure 3). The lowest sensitivity was observed for FLQ (95.1%) due to the presence of heteroresistant *gyr*A populations in each of these isolates which were below the limit of detection in the clinical specimen. Similarly, the A1401G (*rrs*) mutation was identified but at a level below detection of the MTB/XDR assay. The overall sensitivities showed a slight decrease for most drugs and an expected significant decrease for ETH with specificities remaining at >99% against the composite reference standard (Figure 4). Sensitivities of >90% were observed for INH and FLQ, >92% for KAN and CAP, and >96% for AMK which meet the minimum criteria for the target product profile for next generation DST assays. The lower sensitivity of 65.9% (95%CI: 57.1-73.6) observed for ETH compared to 97.6% relative to sequencing, was suggestive of the presence of additional mutations in targets not covered by the assay or false ETH resistance on phenotypic DST. The latter cannot be confirmed since pDST was not performed for discordant results, however ETH pDST is known to be unreliable (22). Based on these findings ETH resistance can be ruled in and not ruled out using the MTB/XDR assay, in agreement with previous observations (23).

Overall, the sensitivities observed for the composite reference were comparable to those previously reported in a geographically diverse setting for INH (90.1% vs 94.0%) and FLQ (90.6% vs 94%) and higher for AMK (96.2% vs 73.0%), KAN (92.7% vs 86.0%), CAP (92.5% vs 61.0%) and ETH (65.9% vs 54.0%) (23). The improved sensitivities highlight the accuracy of the assay in predicting resistance, particularly for INH, FLQ, AMK, KAN and CAP, in our setting. The higher performance observed when using sequencing as a reference is due to evaluation of the regions targeted by the assay only and not considering all resistance conferring mutations across the genome. Further, due to the restricted analysis for sequencing, the discordance observed between phenotypic susceptibility testing and the MTB/XDR test could not be resolved.

The performance of the Xpert XDR in our study for INH resistance detection relative to pDST was similar to the GenoType MTBDRplus 90.7% (95% CI: 84.8 – 94.5) vs 93% (95% CI: 90 – 95) for INH, however, sensitivity for FLQ resistance detection was lower than the GenoType MTBDRsl 90.6% (95%CI: 81.0 – 95.6) vs 95% (95%CI: 91 - 97) (ref 20). When considering performance relative to sequencing, resistance detection by Xpert XDR for INH was superior to the GenoType MTBDRplus 98.5% (95%CI: 94.6 – 99.6) and equivalent to GenoType MTBDRsl for FLQ 95.1 (95%CI: 86.5 – 98.3).

Two shorter standardized regimens for DR-TB are currently recommended by WHO: a regimen for MDR/RR-TB where the injectable agent has been replaced by BDQ (used for 6 months), in combination with levofloxacin (LFX)/moxifloxacin (MXF), ethionamide (ETH), EMB, high-dose INH, PZA and CFZ for 4 months in the intensive phase; and a 6-month regimen for that includes the new anti-TB agent, Pa, which was also recommended by WHO in 2020 for use under operational research conditions, in combination with BDQ and LZD (24). LPAs have been recommended by the WHO for the detection of *Mycobacterium tuberculosis* (*M. tuberculosis*) strains resistant to rifampicin, in both smear positive and smear negative sputum specimens (25, 26). Currently, these LPAs are being used at-scale in South Africa, however, delayed result reporting is a common problem experienced with routine use mainly due to technical aspects such as sample batching. Another challenge with LPAs is its poor sensitivity on smear negative specimens that require repeat testing on cultured isolates. In a recent study, done at two sites in South Africa the median time to LPA results ranged between 5.4 – 23.1 days (27). These delays negatively impact TB control, allowing for the continued transmission of drug resistant *M.* The test is capable of determining level of resistance for both INH and FLQ based on the interpretation of melt-curves detecting the presence of associated mutations. This allows for supporting clinical management by making available the use of both high-dose INH and MFX for patient management, particularly patients requiring a salvage regimen. *tuberculosis* strains and result in inadequate management of patients. Alternative assays providing the same information are still not readily available. Next-generation sequencing-based technologies show early promise, however, none have matured sufficiently to an end-to-end *in vitro* diagnostic assay ready for routine use.

The NHLS is a key partner of the National TB Program and adapts to having the latest and most sensitive diagnostic capability to further strengthen the program. South Africa was the first country to implement the GeneXpert MTB/RIF and GeneXpert MTB/RIF Ultra assay, at scale, as a smear microscopy replacement technology in 2011 and one of the first to perform susceptibility testing for new and repurposed anti-TB drugs (Bedaquiline and Linezolid) as part of diagnosis. Further diagnosis was improved, particularly in HIV co-infected TB-disease individuals, through replacement of GeneXpert MTB/RIF assay with the Xpert MTB/RIF Ultra assay in 2017 with South Africa again taking the lead. These efforts have aided in improved management of people diagnosed with TB and drug-resistant TB, thereby reducing South Africa’s burden of disease.

NHLS has fully implemented this technology following of the WHO recommendation to strengthen the detection and management of drug-resistant TB, an emerging global threat. The deployment of the assay leverages the existing instrument network and testing capacity, increases utilization and thereby maximizes efficiency. The potential benefits are a reduction in public health expenditure, a reduction in the number of repeat tests (through the traditional testing modalities) and a significantly improved turnaround time. Ultimately, turnaround time improvements benefit those individuals’ requiring treatment by enabling clinicians to take earlier appropriate management decisions impacting treatment outcomes.

## Data Availability

All data produced in the present study are available upon reasonable request to the authors

## ACKNOWLEDGEMENTS

We thank the study investigators, clinical research coordinators, and laboratory staff at the Centre for Tuberculosis, NICD/NHLS for their diligent efforts.

The study sponsor (Cepheid) provided the investigational product, funding and administrative and logistical support to this study site. The sponsor was involved in the design, conduct and execution of the clinical study, and assisted in monitoring and collection of data. The sponsor participated in the interpretation of the data and preparation of the manuscript. GL and XL are Cepheid employees.

SVO, LJ, FI, DN, TG, MvdM collected the data and performed the experiments. XL performed the statistical analysis. SVO and GL drafted the manuscript. All authors reviewed and critiqued the draft manuscript and approved the final manuscript prior to submission for publication.

## SUPPLEMENTARY DATA

**Supplementary Table 1:**
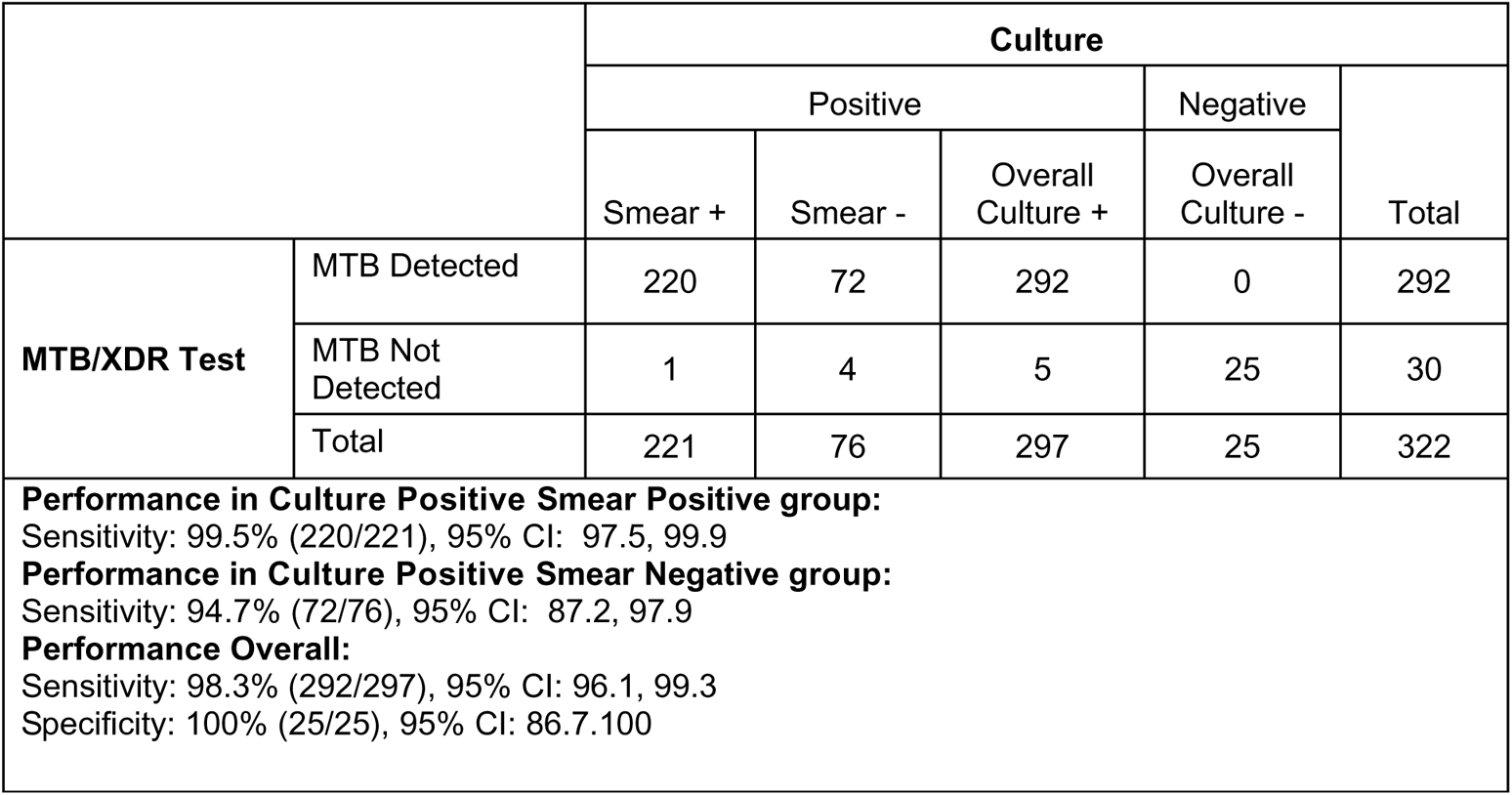
Performance of MTB/XDR test compared to MGIT Culture stratified by smear status.

**Supplementary Table 2:**
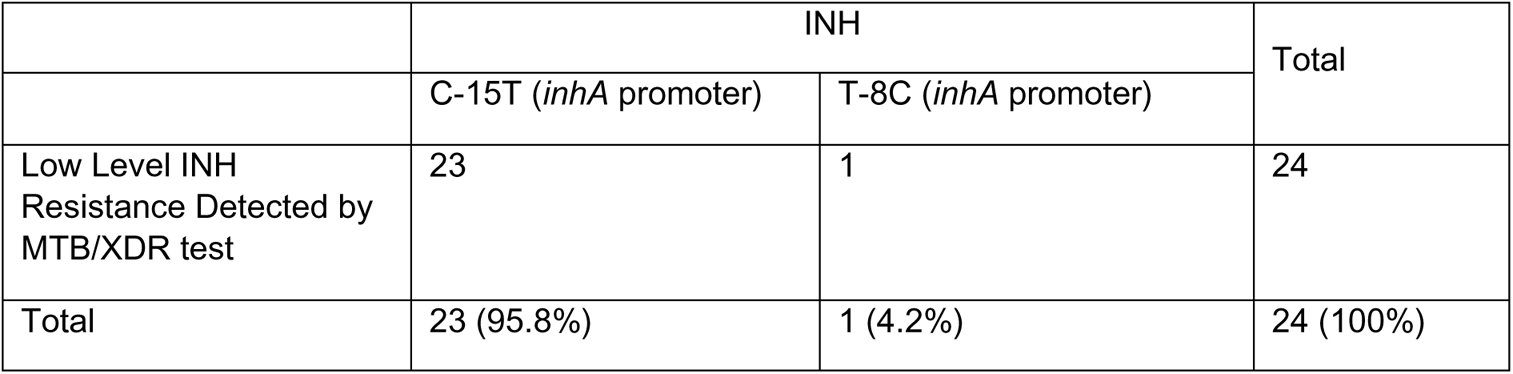
Low Level INH resistance prediction by MTB/XDR test relative to *inhA* promoter mutations identified by sequencing.

**Supplementary Table 3:**
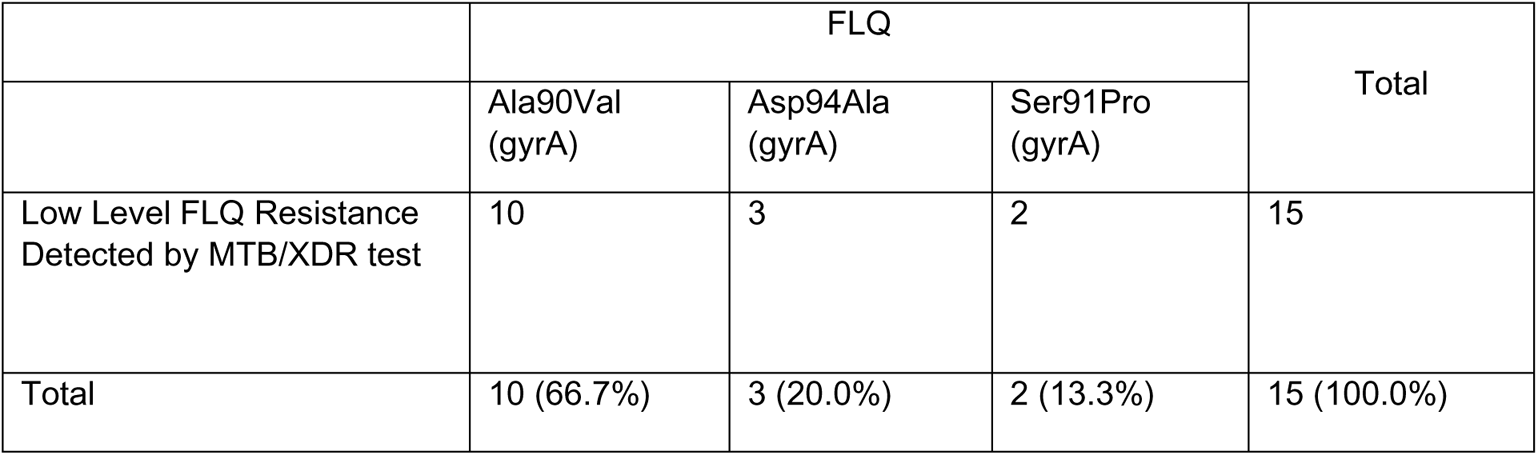
Low level FLQ resistance prediction by MTB/XDR test relative to *gyrA* mutations identified by sequencing.

|  | FLQ |  |  | Total |
| --- | --- | --- | --- | --- |
|  | Ala90Val (*gyrA*) | Asp94Ala (*gyrA*) | Ser91Pro (*gyrA*) |  |
| Low Level FLQ Resistance Detected by MTB/XDR test | 10 | 3 | 2 | 15 |
| Total | 10 (66.7%) | 3 (20.0%) | 2 (13.3%) | 15 (100.0%) |

